# Protocol for assessing the impact of the 6^th^ National Audit Project recommendations on practice

**DOI:** 10.1101/2024.05.03.24306350

**Authors:** Sigrún Eyrúnardóttir Clark, Iain Moppett, S. Ramani Moonesinghe, Cecilia Vindrola-Padros

## Abstract

Patient safety has been a growing area of concern, especially within perioperative care where risks of major complications during surgery exist. Anaphylaxis in the operating theatre is a life-threatening drug reaction that happens suddenly, without warning and can affect anyone. The 6th National Audit Project (NAP6) of the Royal College of Anaesthetists (RCoA): Perioperative Anaphylaxis was the largest ever prospective study of anaphylaxis related to anaesthesia and surgery. The findings from the audit were collated into a report and included recommendations for improved patient care. The purpose of this study is to understand the perceptions of the NAP6 recommendations and their impact on practice. This study will use ethnographic qualitative methods in the form of observations, interviews and a documentary analysis. The sessions targeted for observations will include departmental or hospital meetings, and educational or training sessions, related to perioperative anaphylaxis. The target sample size of 78 healthcare professionals across six hospitals within England, will include individuals with roles specific to anaesthesia, surgery, immunology, allergy and governance. Additionally, six stakeholders will be interviewed who can provide insights into the NAP6 recommendations at the national level. Across the six sites, local collaborators will share any relevant documents related to perioperative anaphylaxis or the NAP-6 recommendations. The study has received regulatory approvals from the Health Research Authority and has been funded by the National Institute for Health and Care Research.

## 2. Introduction

The Royal College of Anaesthetists’ (RCoA) National Audit Projects (NAPs) investigate serious rare complications during anaesthesia and have been recognised as internationally important in terms of their impact on patient outcome [1]. The NAPs have been running for twenty years and have included eight separate audits. The anaesthesia related topics of the audits have ranged from complications of central neuraxial block, complications of airway management, accidental awareness, perioperative anaphylaxis, perioperative cardiac arrest and complications of regional anaesthesia.

The focus of the 6th National Audit Project (NAP-6) was on perioperative anaphylaxis. The audit was conducted across 341 hospitals across the UK, who were asked to share data on the proportion of cases that were referred and investigated; the proportion of cases proven to be anaphylaxis; the consistency in immediate management, referral and investigation with published guidelines; and the correlation between immediate management and outcomes related to perioperative anaphylaxis. The findings from the audit were collated into the 2018 report by the RCoA in collaboration with the National Institute of Academic Anaesthesia, Health Services Research Centre and the National Audit Project [2]. In addition to summarising the findings, the report also shared recommendations for quality enhancements and improved patient care in the field of perioperative anaphylaxis.

Patient safety has been a growing area of concern within healthcare, as global estimates have shown that annually within healthcare there are more than 3 million deaths due to unsafe care, and around 1 in every 10 patients faces harm [3]. A key setting where patient safety is of even more concern is within perioperative care, due largely to the risk of major complications during surgery with the need for rapid recognition and action; and the context of the health of the UK with many patients risk to surgery being compounded by long-term conditions such as diabetes and heart disease [4,5]. In response to this context the Central London Patient Safety Research Collaboration (PSRC) was formed to deliver world-class research into improving the safety of Surgical, Perioperative, Acute and Critical care services (SPACE).

The particular focus of this study protocol is on the recommendations made by the RCoA – with an ethnographic investigation of the perceptions and experiences of healthcare staff and national bodies in response to the NAP6 recommendations. Which fits within the Learning cluster of the PSRC with the focus on improving organisational patient safety culture and practice. The purpose of the study is to understand perceptions of NAP6 recommendations, if any of these recommendations are being used in practice, any factors acting as barriers or enablers in the implementation of the recommendations, and if any improvements can be made to future NAP recommendations to increase their use in practice.

### Aims and objectives

1. Investigate NHS healthcare staff and national body members’ perceptions of the NAP6 recommendations.
2. Assess which recommendations were translated into changes in practice.
3. Describe the measured or assumed impact of the implementation of recommendations.
4. Identify factors that acted as enablers in the translation of the recommendations into changes in practice.
5. Identify factors that acted as barriers in the translation of the recommendations into changes in practice.
6. Explore if any lessons can be suggested for the development of future NAP recommendations.

## 3. Methods

This study will use ethnographic qualitative methods in the form of observations, interviews and a documentary analysis.

### 3.1. Sampling approach

Several NAP Local Coordinators (LCs) who were appointed across England will be selected using convenience sampling based on existing connections within the research team, to ask if they or their colleagues would take on the role as a local investigator and involve their hospital(s) in the study.

#### NHS hospital sites

Between four to six different hospitals will be recruited to the study using purposive sampling to identify accessible sites that demonstrate diversity in terms of their geographical location and NHS provider sector (secondary or tertiary care hospitals).

#### Participants

The local investigator at each site will be asked to identify suitable sessions to observe, and to identify and approach potential participants for interviews. The desired sample is 10-13 stakeholders per site, which may lead to an overall sample size of 78 interviews. Additionally, stakeholders will be identified by the research team, who can provide insights into the NAP6 recommendations at the national level. Potential interviewees will be purposively selected to capture data from the following job roles:

1. Micro level: Primary perioperative anaphylaxis resuscitation providers
  - Anaesthetist 1 (e.g., team leader)
  - Anaesthetist 2 (e.g., trainee)
  - Anaesthesia Associate
2. Meso level: Perioperative anaphylaxis resuscitation team
  - Anaesthetist 3 (e.g., called to assist)
  - Surgeon
  - Operating department practitioner
  - Theatre nurse
  - Intensive care staff (intensivist, critical care nurse)
  - Other (may be nominated by interviewees)
3. Macro level: Anaesthetic department and wider Trust organisation
  - Resuscitation lead (anaesthetist)
  - Department lead (anaesthetist)
  - Governance/safety lead (anaesthetist)
  - Departmental anaphylaxis lead
  - Immunology lead
  - Microbiology lead
  - Allergy clinic staff (consultant allergist)
  - Clinical or medical director
  - Theatre managers
  - Trust safety lead
  - Emergency planning lead
  - Training lead
  - Other (may be nominated by interviewees)
4. National:
  - Members of the RCoA
  - Members of the British Society for Allergy and Clinical Immunology (BSACI)

#### Documents

Relevant documents for analysis will be purposively selected and shared by local investigators, documents will be related to perioperative anaphylaxis and/or NAP-6.

### 3.2. Data collection

#### Observations

##### Aim relevant to this form of data collection

Assess if any recommendations were translated into changes in practice and if measured, the impact of the implementation of recommendations. The relevant sessions identified by NAP local co-ordinators for non-participatory observation could include formal departmental or hospital meetings or informal discussions; or educational or training sessions, related to perioperative anaphylaxis. An observation guide will be used to collected standardised data across sites.

#### Interviews

##### The interviews will aim to explore the following topics

- Staff perceptions of the NAP6 recommendations.
- Recommendations that have been translated into changes in practice at the individual, team and organisational level.
- Measured or assumed impact of the implementation of recommendations will also be assessed.
- Factors that have enabled the translation of recommendations into changes in practice.
- Factors that have hindered the translation of recommendations into changes in practice.
- Recommendations of improvements to the future development of NAP recommendations.

Semi-structured interviews with healthcare professionals and members of national bodies will be conducted using semi-structured topic guides, which may undergo adaptations based on emerging findings from the interviews. There will be no financial incentive for participation, so interviews will be voluntary and conducted on a one-to-one basis. The contents of the interviews will be audio-recorded and transcribed.

#### Documentary analysis

##### Aim relevant to this form of data collection

Assess if any recommendations were translated into changes in practice and if measured, the impact of the implementation of recommendations. Localised and public documents related to the training for, and management of perioperative anaphylaxis will be reviewed. A documentary analysis form will used to capture standardised findings across the documents.

### 3.3. Data analysis

A deductive approach will initially take place as interview data collection and analysis is ongoing, any key themes related to the interview topic guide will be inserted into RREAL rapid assessment procedure (RAP) sheets [6]. An inductive thematic approach will also be applied as new data patterns emerge and these will be inserted into the RREAL RAP sheets. The findings from the RREAL sheet will support with sharing project updates, updating topic guides and the development of the codebook for the in-depth analysis. The codebook will be validated by two researchers coding a sub-sample of the data sources.

During the in-depth framework analysis [7], NVivo10 will be used to triangulate and analyse the imported interview transcripts, documents and observation field notes. Data will be deductively coded into themes that address the research question, and inductively into any new emerging themes from the data. The team will meet iteratively to discuss any significant findings and queries from the coding process.

### 3.4. Regulatory approvals

This study has been funded by the National Institute for Health and Care Research and has received regulatory approvals from the Health Research Authority. Across all interviews, stakeholders will receive a participant information sheet and be given time to review the contents. All interviewees will then share written consent prior to participating in any interview. Prior to the observations another participant information sheet will be circulated with all staff who may be attending the session, should any participant wish to opt out of the observation the researchers will not record the participants input within their field notes.

## Data Availability

All data produced in the present study are available upon reasonable request to the authors.

